# ORIGINAL RESEARCH; QUALITY ASSESSMENT AND COMPARATIVE ANALYSIS ON THE RECOMMENDATIONS OF CURRENT GUIDELINES ON SCREENING AND DIAGNOSIS OF PERIPHERAL ARTERIAL DISEASE; A SYSTEMATIC REVIEW

**DOI:** 10.1101/2022.02.02.22270053

**Authors:** O Uyagu, C Ofoegbu, J Ikhidero, E Chukwuka, O Enwere, O Ogierhiakhi, A. Adelosoye

## Abstract

**Introduction:** Peripheral Artery Disease (PAD) is a major atherosclerotic disease, and there are several clinical practice guidelines available for it. The paucity of strong evidence is known to give room for variations in recommendations across guidelines with attendant confusion amongst clinicians in clinical practice. This study aims to conduct a quality assessment and comparative analysis on PAD screening and diagnostic recommendations in the management of PAD.

**Methods:** We conducted a systematic review of CPGs’ written after 2010 and on or before 2020. An exhaustive search was conducted through the major medical databases and websites of specialist international organisations of interest and using our inclusion criteria, the appropriate guidelines were extracted. The AGREE-II instrument was used for quality assessment, while the recommendations across screening and diagnosis were extracted and then comparatively analysed.

**Results:** We found nine guidelines that fit our criteria. The guidelines had the lowest scores across the applicability and stakeholder involvement domains. The highest scores were recorded in the Clarity of presentation, Scope and purpose and Editorial independence in order of decreasing magnitude. Also, the trend was the guideline quality scores improved over time. The guidelines were unanimous in offering to screen to ‘high-risk ‘patients, although there were some discrepancies in the appropriate age range and unavailability of strong evidence across the guidelines backing this recommendation. The guidelines also showed harmony in adopting the Ankle-Brachial index as the initial diagnostic investigation of choice. However, concerning further diagnostic investigations and imaging, we found several discrepancies among the recommendations in the absence of strong evidence.

**Conclusion:** Though the quality of the guidelines is shown to be improving over time, they display poor scores in the stakeholder involvement and applicability domains, which could be influencing low interest in research that can improve screening and diagnostic recommendations.

**STRENGHTS AND LIMITATIONS:**

- This review, unlike previous studies, focused on Peripheral Arterial Diseases(PAD) guidelines written after 2010 and reflects a synthesis of the current state of guideline quality and the most recent recommendations in PAD management regarding screening and diagnosis.
- Complex data has been aggregated, comparatively assessed using thematic analysis and the results presented in concise and straightforward forms using texts, charts and tables to satisfy the needs of all kinds of readers alike from the medical research community to the patients and public reader.
- By utilising rigorous systematic review methodology and a mixed qualitative and quantitative approach to the data analysis, this study has revealed the current areas of strengths and weaknesses of the quality of the PAD guidelines, which is inadvertently related to the reason behind the persisting absence of high-level evidence in screening and diagnostic recommendations.
- Qualitative analyses are inherently challenging to process, especially when dealing with clinical practise guidelines (CPGs’) that contain large amounts of information; as such, the process was cumbersome and time-consuming with the inevitable loss of data during the thematic classification process.
- During the literature search, the search strategies were executed exclusively in English Language labouring under the auspices that the major PAD CPG’s will have an English language translation, so it is possible that some guidelines written within the study timeframe were not captured due to this limitation.

**Registration:** Registrated in PROSPERO; ID; CRD42020219176

## INTRODUCTION

Atherosclerotic disease is an umbrella term for the world’s leading cause of mortality and morbidity. (1) Peripheral artery disease (PAD) is a major component of this group of disorders after cerebrovascular and coronary artery disease, sharing the same risk factors with the other atherosclerotic conditions. (2) Interestingly, according to data from the REACH registry, it was observed that individuals with PAD. do not achieve risk factor control as frequently as those with Coronary Artery Disease (CAD) and Cerebrovascular Disease (CVD), and in addition, had higher levels of mortality comparatively. (3) The apparent explanation for this is that PAD is the most under-diagnosed and poorly treated atherosclerotic disease. PAD is a chronic medical disease with an asymptomatic phase of variable duration, with some individuals progressing into the symptomatic phase. Optimal management mainly involves early identification of the condition (screening and diagnosis), optimal medical management, which requires risk factor modification (through pharmacological and non-pharmacological methods), supervised exercise therapy and sometimes revascularisation.

Clinical Practice Guidelines (CPGs) have methodically developed statements to guide physicians and patients in making safe healthcare decisions based on the best available evidence. (4,5) Currently, there are some CPGs outlining best practices in the management of PAD. The quality of the CPGs varies between the authoring organisations and is also influenced by time as new evidence comes to light, ushering changes to guideline recommendations. As such, systematic reviews on the guidelines of particular disorders are often conducted; this study will review the quality of the guidelines available on PAD and assess the variations in their recommendations with regards to the core aspects of management. A few partial reviews have been conducted on aspects of Peripheral Artery Disease (PAD) guidelines in the past (6–8). Our study encompasses all aspects of PAD. Management from screening and diagnosis, through medical management, to revascularisation and follow up. Due to the volume of findings, the paper has been split into three papers, with this being the first of the series. This paper encompasses the quality assessment and critical analysis of recommendations across screening and diagnostic recommendations. Also, we have limited the publication date range for the CPGs from after the year 2010 until 2020 to get the most recent information on PAD. Management recommendations, unlike the previous reviews which scanned guidelines over a wide range of time. As such, the risk of evaluating outdated information is avoided.

As outlined in our published protocol, (9) this paper aims to elucidate with diligent analysis, evaluation and crisp data presentation the quality of the current guidelines on PAD, with recommendations on their suitability for use in clinical practice. In addition, we intend to review the long-standing debate on screening and diagnostic recommendations to ascertain the level of variation between authoring organisations. We expect that there should be greater levels of harmony with new evidence compared to older guideline reviews. Also, areas of interest where recommendations vary due to low-level evidence will be elucidated.

## MATERIALS AND METHODS

A systematic search was conducted, and eligible guidelines were selected based on the attributes listed in the PICAR (Population, Intervention, Comparator, Attribute, Recommendation Characteristics) statement of our published protocol. (9) The Preferred Reporting Items for Systematic Reviews and Meta-Analysis statement was used as a reference to report items and results in this study. (10)

### Patient and public involvement statement

Patients who are members of the Peripheral Arterial Diseases Support Group (https://www.facebook.com/groups/pad.pvd.support/members) were involved in this study’s design (in modelling the research objectives). The Way to My Heart.org (https://www.thewaytomyheart.org/) founded this support group. The patient public involvement is coordinated through the group’s leaders/founders (also patients themselves are actively involved in providing support to their fellow patients) who are advisory members to the research team. They have identified this research as a priority area for clinicians who provide care to patients living with PAD. The group members have been informed of this study’s results through their leadership. The support group will also participate in publicising the study after publication.

### Search Strategy

A systematic search was performed to identify relevant CPGs on PAD. One reviewer (O.U) conducted the search and extraction in line with the inclusion and exclusion criteria and this was independently verified by a second reviewer (C. O). A third reviewer (J.I) was called in to resolve differing results. We developed a concept table to generate appropriate search terms (MeSH, Free text vocabulary, Key Words) depending on the database’s peculiarities. Databases searched included Scopus (which includes Embase and Medline), TRIP and Cochrane. The search also included guideline developer websites such as NICE, SIGN, NIH, GIN and websites for national academic societies. Details of the search strategies can be found in Appendix 3 below and the protocol.

### Selection of Guidelines

In line with our protocol, guidelines that met the following inclusion criteria were selected.

1. The guideline is a CPG developed for people with PAD.
2. The guideline covers recommendations regarding screening, non-pharmacological and pharmacological interventions, surgical and follow-up management.
3. The guidelines were written after 2010 and in or before 2020.
4. The guideline is the most recent version.
5. The guideline is available online.
6. Related or international academic organisations wrote the guideline.

Our exclusion criteria were.

1. The topic is only mentioned in the guideline.
2. The guideline is limited to a specific aspect of PAD. Management, such as screening, pharmacologic management, etc.

#### Outcomes

The primary outcome sought in this study were; Guideline Quality and Guideline recommendations on screening and diagnostic methods. Secondary outcome data included guideline characteristics; year of writing, funding source, language of writing, location, website/source.

### Quality Assessment

In this study, the updated AGREE-II instrument was used to assess the quality of the selected guidelines. The AGREE-II instrument is a 23-item tool with international certification that evaluates the six methodological quality domains of a guideline, including scope and purpose, stakeholder involvement, the rigour of development, clarity of presentation and applicability and editorial independence. (11) As was written in the protocol, the assessment was conducted by four reviewers (as recommended by the tool’s developers to minimise bias) using the instrument to assess all selected guidelines. The reviewers scored each guideline across each domain on a Likert scale of 1 through 7 (from strongly disagree to strongly agree). In addition, the reviewers gave an overall score of the guidelines on a similar Likert scale. As such, each guideline has two sets of scores: (a) the domain scores and (b) the overall score for the guideline. The details for the scoring system of the AGREE instrument is outlined in the protocol (9).

The overall quality assessment was arrived at using the domain scores in line with the study protocol. Guidelines with four or more domains scored over 60%, would be regarded as ‘strongly recommended for use in practice’; if scores of most domains (four or more) ranged 30%–60%, the guideline was considered ‘recommended for use with some modification’. Those with domain scores (four or more) less than 30% were regarded as ‘not recommended for use in practice’. The overall guideline scores were used as a supporting statistic only and did not directly contribute to the grading of guideline quality. The data set for the AGREE-II scores was submitted in a public data repository accessible at http://www.doi.org/10.11922/sciencedb.01479.

### Guideline Recommendations

The recommendations were extracted into a matrix in Microsoft Excel sheets. Then thematic analysis was utilised to organise the recommendations into themes which allowed us to summarise the information into tables for comparisons. The strength of recommendations and level of evidence was extracted and displayed in the tables for each recommendation. Each guideline used its grading method, which we harmonised using our grading system for the purpose of comparison for this study (Table 1 & 2below)

**Table 1.**
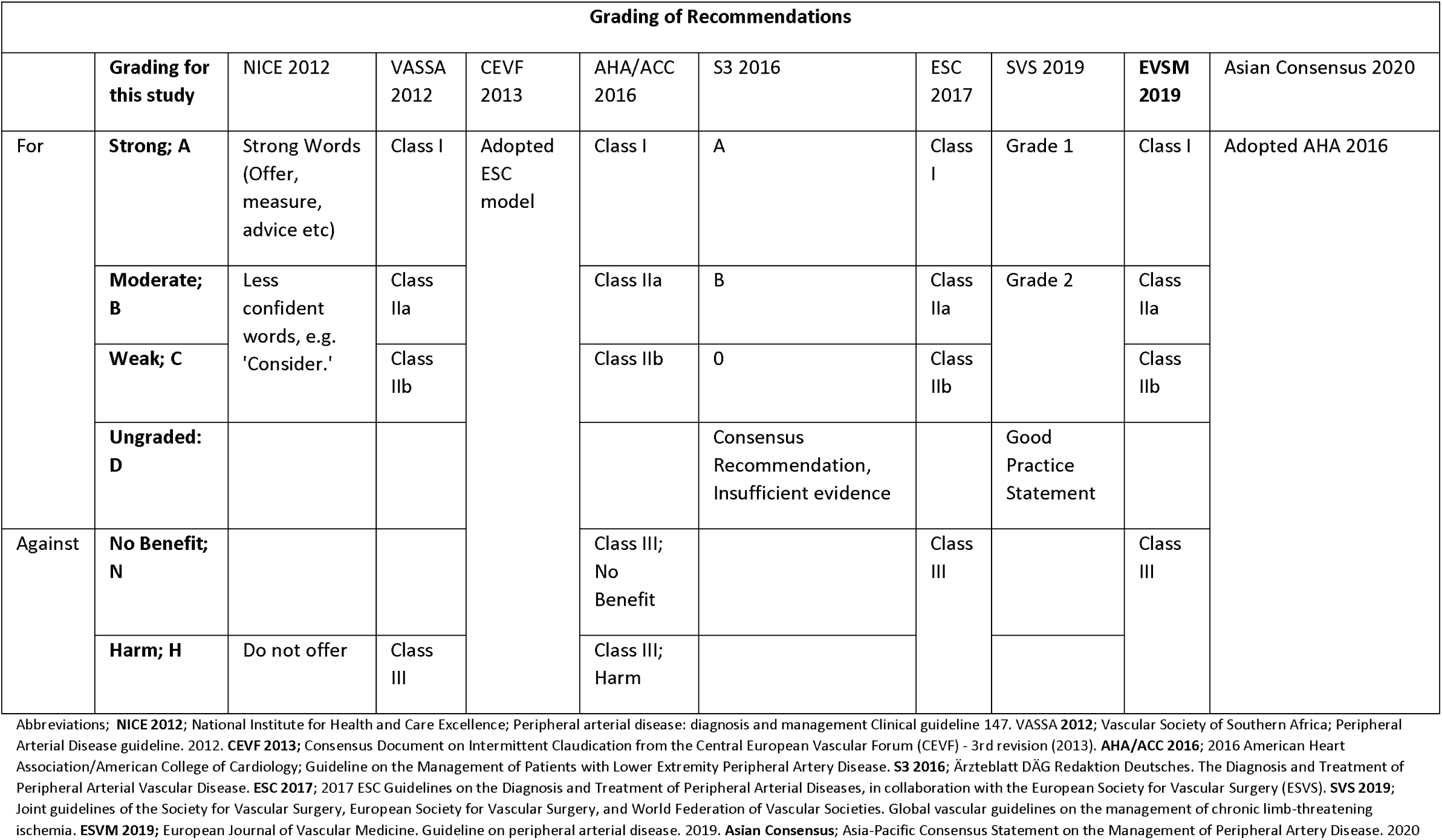
Harmonizing Recommendation strength grading system across the guidelines.

**Table 2.**
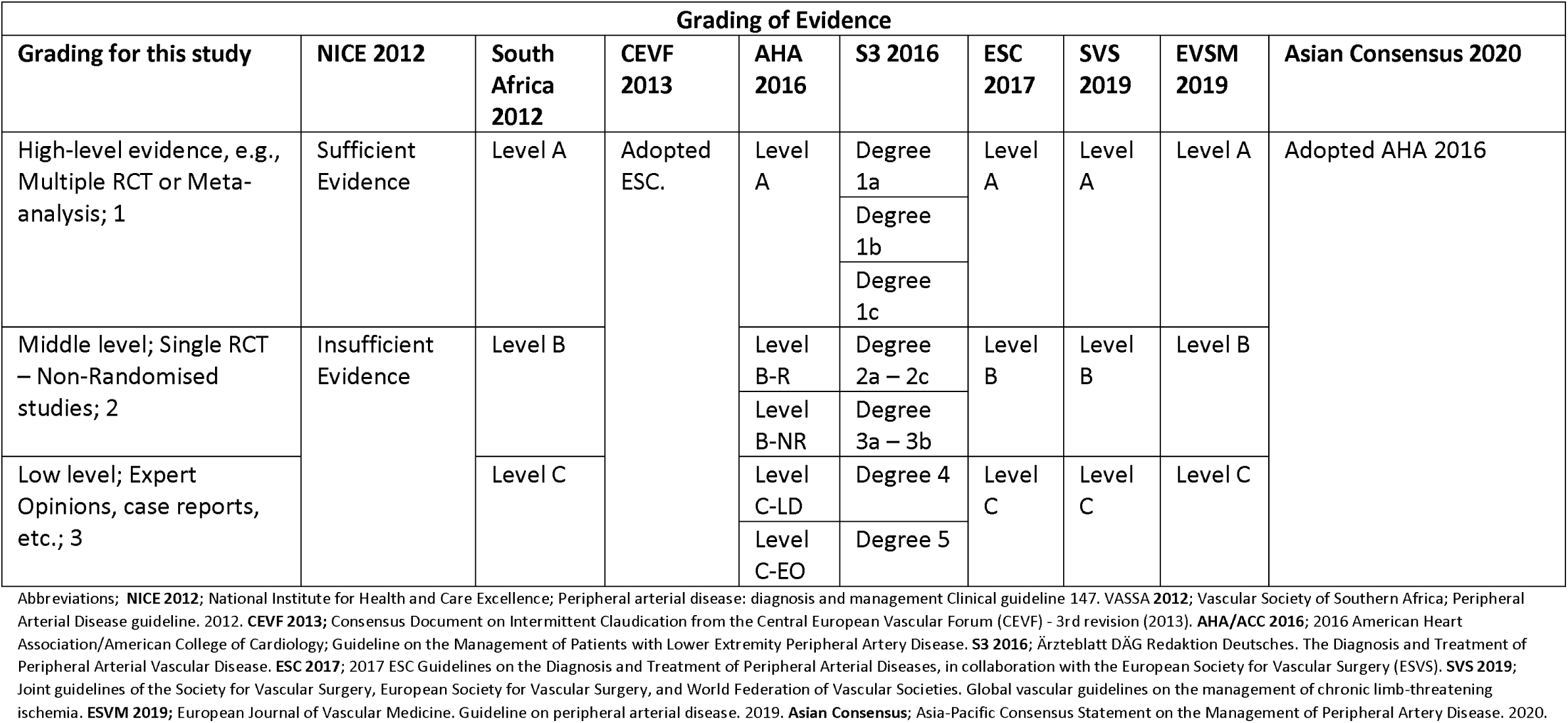
Harmonizing Level of Evidence grading system across the guidelines.

| Grading of Evidence |  |  |  |  |  |  |  |  |  |
| --- | --- | --- | --- | --- | --- | --- | --- | --- | --- |
| Grading for this study | NICE 2012 | South Africa 2012 | CEVF 2013 | AHA 2016 | S3 2016 | ESC 2017 | SVS 2019 | EVSM 2019 | Asian Consensus 2020 |
| High-level evidence, e.g., Multiple RCT or Meta-analysis; 1 | Sufficient Evidence | Level A | Adopted ESC. | Level A | Degree 1a | Level A | Level A | Level A | Adopted AHA 2016 |
|  |  |  |  |  | Degree 1b |  |  |  |  |
|  |  |  |  |  | Degree 1c |  |  |  |  |
| Middle level; Single RCT – Non-Randomised studies; 2 | Insufficient Evidence | Level B |  | Level B-R | Degree 2a – 2c | Level B | Level B | Level B |  |
|  |  |  |  | Level B-NR | Degree 3a – 3b |  |  |  |  |
| Low level; Expert Opinions, case reports, etc.; 3 |  | Level C |  | Level C-LD | Degree 4 | Level C | Level C | Level C |  |
|  |  |  |  | Level C-EO | Degree 5 |  |  |  |  |

Abbreviations; **NICE 2012**; National Institute for Health and Care Excellence; Peripheral arterial disease: diagnosis and management Clinical guideline 147. **VASSA 2012**; Vascular Society of Southern Africa; Peripheral Arterial Disease guideline. 2012. **CEVF 2013**; Consensus Document on Intermittent Claudication from the Central European Vascular Forum (CEVF) - 3rd revision (2013). **AHA/ACC 2016**; 2016 American Heart Association/American College of Cardiology; Guideline on the Management of Patients with Lower Extremity Peripheral Artery Disease. **S3 2016**; Ärzteblatt DÄG Redaktion Deutsches. The Diagnosis and Treatment of Peripheral Arterial Vascular Disease. **ESC 2017**; 2017 ESC Guidelines on the Diagnosis and Treatment of Peripheral Arterial Diseases, in collaboration with the European Society for Vascular Surgery (ESVS). **SVS 2019**; Joint guidelines of the Society for Vascular Surgery, European Society for Vascular Surgery, and World Federation of Vascular Societies. Global vascular guidelines on the management of chronic limb-threatening ischemia. **EVSM 2019**; European Journal of Vascular Medicine. Guideline on peripheral arterial disease. 2019. **Asian Consensus**; Asia-Pacific Consensus Statement on the Management of Peripheral Artery Disease. 2020.

One reviewer performed extractions and then reviewed for completeness and consistency by another reviewer, after which comparisons were made across the guidelines.

## RESULTS

### Search Results

The initial search identified three thousand one hundred and forty-nine citations. The flowchart (Figure 1) shows how we systematically eliminated the guidelines by removing duplicates, previous versions and guidelines written outside the date range, screening the title and abstracts for citations not related to the topic, eliminating those not CPGs, and finally eliminating those which targeted aspects of PAD. Management of special populations. In the end, we had 9 CPGs, which were included in this study for analysis (12–20).

**Figure 1.**
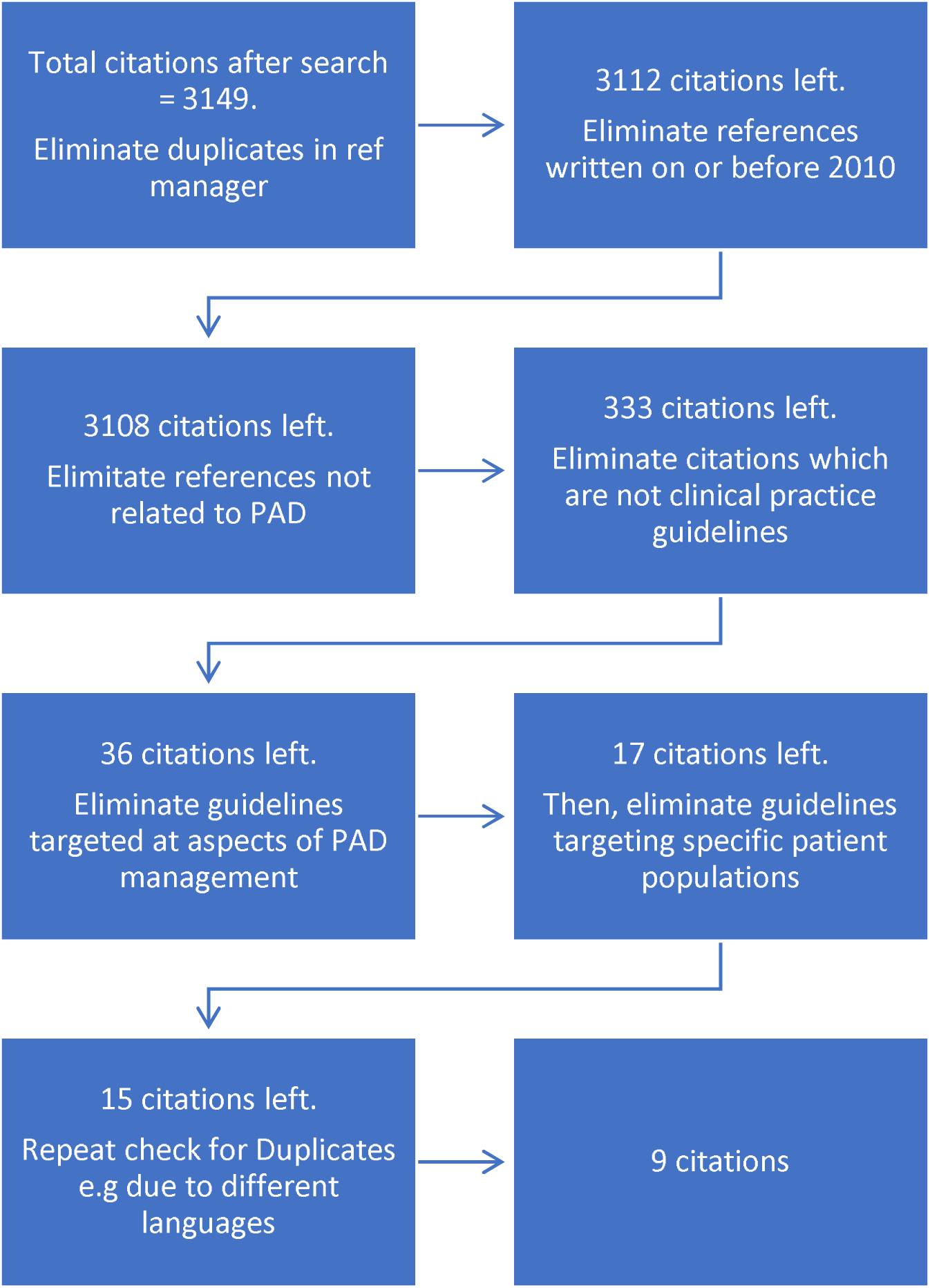
Flow chart of the search strategy.

### Guideline characteristics

The guidelines included are presented in Table 3 below. They were written after 2010 and before or in 2020.The majority of the guidelines (eight) were written in English, except the German guideline, which was written in German. The extended German guideline was translated into English for analysis, while a short version was already translated to English. Two guidelines did not state their source of funding (VASSA and CEVF). The overall AGREE score on guideline quality ranged from 68 to 84.

**Table 3.** Characteristics of Included Guidelines.

| CPG | Developing Organization | Country | Language of Publication | Date Of Search | Date Of release | Publication Site | Funding | Overall AGREE score |
| --- | --- | --- | --- | --- | --- | --- | --- | --- |
| NICE 2012 | National Health System | UK | English | 2020 | 2012 | https://www.nice.org.uk/guidance/cg147 | NHS | 71 |
| VASSA 2012 | One academic society | South Africa | English | 2020 | 2012 | http://www.vascularsociety.co.za/wp-content/uploads/2015/08/Peripheral-Arterial-Disease-VASSA-practice-guidelines-2012.pdf | Not Stated | 68 |
| CEVF 2013 | One academic society | Europe | English | 2020 | 2013 | https://www.minervamedica.it/en/journals/international-angiology/article.php?cod=R34Y2014N04A0329 | Not stated | 68 |
| S3 2016 | One academic society | Germany | German | 2020 | 2016 | https://www.aerzteblatt.de/int/archive/article/183158/The-diagnosis-and-treatment-of-peripheral-arterial-vascular-disease | German Society for Angiology | 82 |
| AHA/ACC 2016 | Two Academic societies | USA | English | 2020 | 2016 | https://www.sciencedirect.com/science/article/pii/S0735109716369029?via%3Dihub | No commercial sponsor | 83 |
| ESC 2017 | Two Academic Societies | Europe | English | 2020 | 2017 | https://academic.oup.com/eurheartj/article/39/9/763/4095038 | No commercial sponsor | 82 |
| SVS 2019 | 3 Academic Societies | Global | English | 2020 | 2019 | https://linkinghub.elsevier.com/retrieve/pii/S0741521419303210 | No commercial sponsor | 82 |
| ESVM 2019 | 1 Society | Europe | English | 2020 | 2019 | https://econtent.hogrefe.com/doi/full/10.1024/0301-1526/a000834?rfr_dat=cr_pub++0pubmed&url_ver=Z39.88-2003&rfr_id=ori%3Arid%3Aacrossref.org | No external sponsor | 86 |
| Asian Consensus 2020 |  | Asia | English | 2020 | 2020 | https://www.jstage.jst.go.jp/article/jat/27/8/27_53660/article | No external sponsor | 86 |

Abbreviations; CPG; Clinical Practice Guideline. NICE 2012; National Institute for Health and Care Excellence; Peripheral arterial disease: diagnosis and management Clinical guideline 147. VASSA 2012; Vascular Society of Southern Africa; Peripheral Arterial Disease guideline. 2012. CEVF 2013; Consensus Document on Intermittent Claudication from the Central European Vascular Forum (CEVF) - 3rd revision (2013). AHA/ACC 2016; 2016 American Heart Association/American College of Cardiology; Guideline on the Management of Patients With Lower Extremity Peripheral Artery Disease. S3 2016; Ärzteblatt DÄG Redaktion Deutsches. The Diagnosis and Treatment of Peripheral Arterial Vascular Disease. ESC 2017; 2017 ESC Guidelines on the Diagnosis and Treatment of Peripheral Arterial Diseases, in collaboration with the European Society for Vascular Surgery (ESVS). SVS 2019; Joint guidelines of the Society for Vascular Surgery, European Society for Vascular Surgery, and World Federation of Vascular Societies. Global vascular guidelines on the management of chronic limb-threatening ischemia. ESVM 2019; European Journal of Vascular Medicine. Guideline on peripheral arterial disease. 2019. Asian Consensus; Asia-Pacific Consensus Statement on the Management of Peripheral Artery Disease. 2020. UK; United Kingdom. USA; United States of America

### Guideline Appraisal

The standardised scores for each guideline were calculated according to the formula provided by the AGREE tool developers (9). The scores were displayed with a radar chart which allowed for easy comparison of all the guidelines included in this study across domains in Figure 2 below. To give a general overview for the domains, Scope and Purpose; Range 60 – 90, with a mean(SD) of 78.4(11.4), Stakeholder Involvement; Range 50 – 88, with a mean(SD) of 65.3 (13), Rigor of Development; Range 43 – 82, with a mean(SD) of 70 (11.7), Clarity; Range 75 – 94, with a mean(SD) of 86.8 (5.1), Applicability; Range 46 – 77 with a mean(SD) of 62 (9.9), Editorial Independence; Range 44 – 94 with a mean(SD) of 76.2 (18.6) and Overall quality; Range 68 – 86 with a mean of 78.5 (7.2). The domains with the highest score were Clarity of presentation, Scope and purpose and Editorial independence in order of decreasing magnitude. In contrast, Applicability and Stakeholder Involvement tied domains with the lowest scores. Seven guidelines met the criteria for high-quality guidelines, while two, the CEVF and South African guidelines, were recommended for use with some modification as moderate quality guidelines.

**Figure 2.**
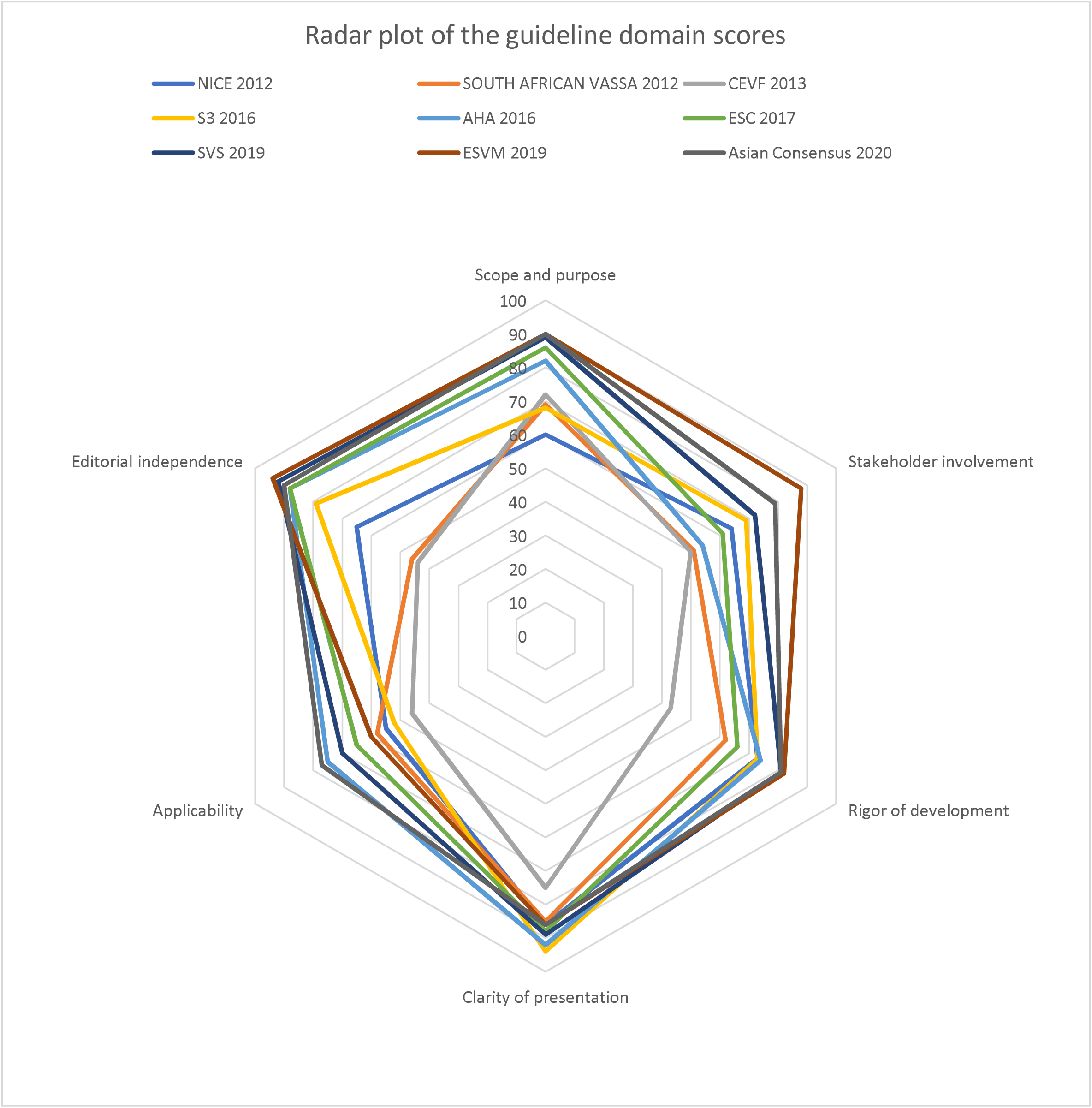
Radar chart showing the domain scores of the included guidelines. Abbreviations; **NICE 2012**; National Institute for Health and Care Excellence; Peripheral arterial disease: diagnosis and management Clinical guideline 147. VASSA **2012**; Vascular Society of Southern Africa; Peripheral Arterial Disease guideline. 2012. **CEVF 2013**; Consensus Document on Intermittent Claudication from the Central European Vascular Forum (CEVF) - 3rd revision (2013). **AHA/ACC 2016**; 2016 American Heart Association/American College of Cardiology; Guideline on the Management of Patients with Lower Extremity Peripheral Artery Disease. **S3 2016**; Ärzteblatt DÄG Redaktion Deutsches. The Diagnosis and Treatment of Peripheral Arterial Vascular Disease. **ESC 2017**; 2017 ESC Guidelines on the Diagnosis and Treatment of Peripheral Arterial Diseases, in collaboration with the European Society for Vascular Surgery (ESVS). **SVS 2019**; Joint guidelines of the Society for Vascular Surgery, European Society for Vascular Surgery, and World Federation of Vascular Societies. Global vascular guidelines on the management of chronic limb-threatening ischemia. **ESVM 2019**; European Journal of Vascular Medicine. Guideline on peripheral arterial disease. 2019. **Asian Consensus**; Asia-Pacific Consensus Statement on the Management of Peripheral Artery Disease. 2020.

Another area of interest was to see the performance of the guidelines over time. The line chart below (Figure 3) showed the composite scores across domains for each guideline plotted over time. We can see clearly; the general trend shows the guidelines increasing in quality from 2012 through 2020.

**Figure 3.**
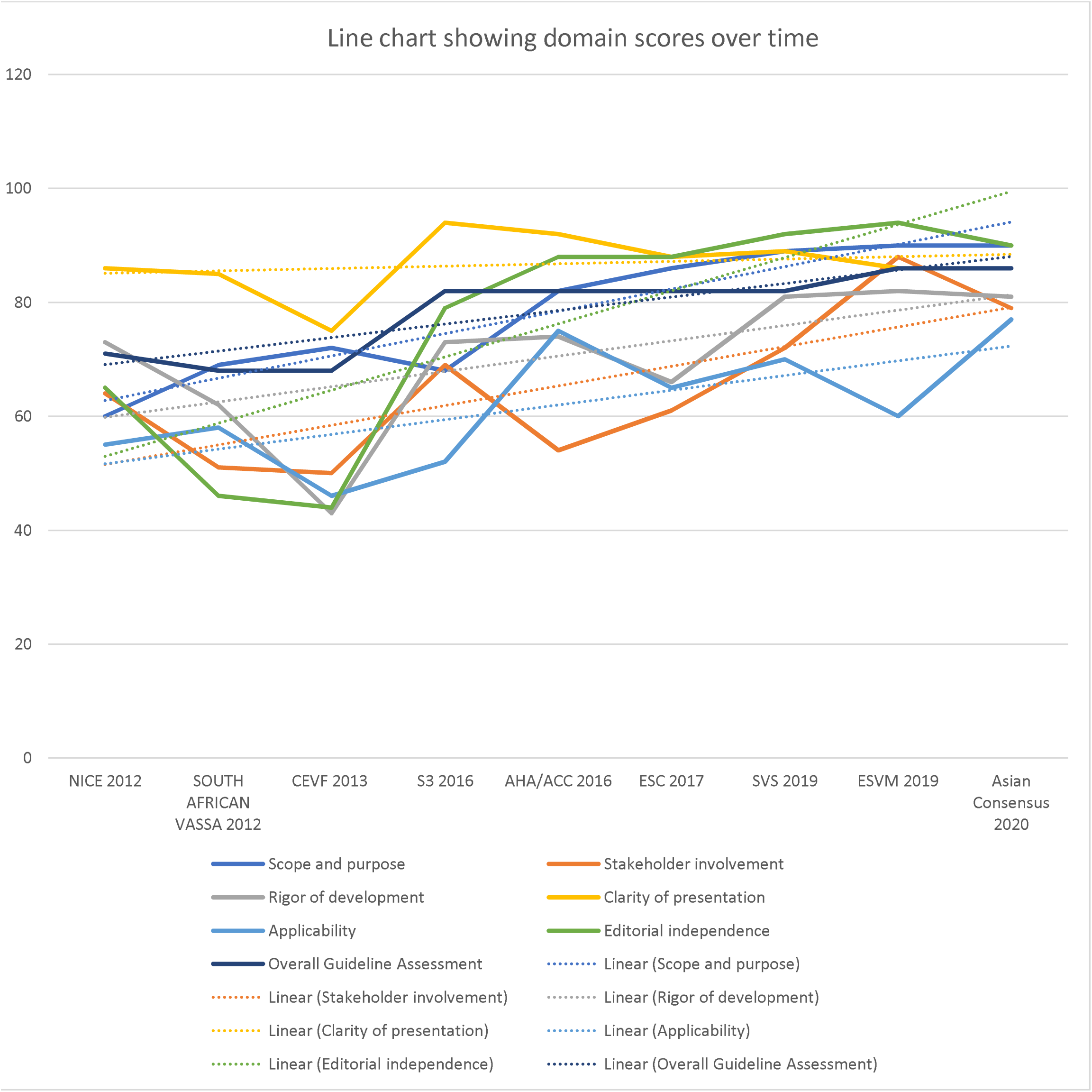
Time trend chart for the domain scores of the included guidelines. Abbreviations; **NICE 2012**; National Institute for Health and Care Excellence; Peripheral arterial disease: diagnosis and management Clinical guideline 147. VASSA **2012**; Vascular Society of Southern Africa; Peripheral Arterial Disease guideline. 2012. **CEVF 2013**; Consensus Document on Intermittent Claudication from the Central European Vascular Forum (CEVF) - 3rd revision (2013). **AHA/ACC 2016**; 2016 American Heart Association/American College of Cardiology; Guideline on the Management of Patients with Lower Extremity Peripheral Artery Disease. **S3 2016**; Ärzteblatt DÄG Redaktion Deutsches. The Diagnosis and Treatment of Peripheral Arterial Vascular Disease. **ESC 2017**; 2017 ESC Guidelines on the Diagnosis and Treatment of Peripheral Arterial Diseases, in collaboration with the European Society for Vascular Surgery (ESVS). **SVS 2019**; Joint guidelines of the Society for Vascular Surgery, European Society for Vascular Surgery, and World Federation of Vascular Societies. Global vascular guidelines on the management of chronic limb-threatening ischemia. **ESVM 2019**; European Journal of Vascular Medicine. Guideline on peripheral arterial disease. 2019. **Asian Consensus**; Asia-Pacific Consensus Statement on the Management of Peripheral Artery Disease. 2020.

### Guideline Recommendations

#### Screening Recommendations

All included guidelines unanimously recommend screening high-risk groups, as seen in Table 4 above. Recommendations against screening groups not at risk were given by the ACC/AHA guideline and the Asian Consensus. The strength of recommendations was predominantly strong (except for the AHA guideline and Asian Consensus Statement). The evidence levels for this recommendation were predominantly moderate except for the German S3 guideline, which relied on strong evidence and ESC, which utilised weak evidence.

**Table 4.** Summary of the Screening recommendations for the included guidelines.

| CPG. | Recommendation | Strenght | Evidence | Target Population | Screening Test | Further Testing | Intervals | Intervention for High-Risk groups |
| --- | --- | --- | --- | --- | --- | --- | --- | --- |
| **NICE 2012** | NR. | - | - | - | - |  |  |  |
| **VASSA 2012** | For | A | 2 | Increased Risk* | ABI | Recommended** |  | Recommended** |
| **CEVF 2013** | For | A | 2 | Increased Risk* | ABI | Recommended** | 2-3Years | Recommended** |
| **S3 2016** | For | A | 1 | Increased Risk | ABI |  |  |  |
| **AHA/ACC 2016** | For | B | 2 | Increased Risk | A.B.I. |  |  | Recommended** |
|  | Against | N | 2 | No risk |  |  |  |  |
| **ESC 2017** | For | A | 3 | Increased Risk | ABI. |  |  | Recommended** |
| **SVS 2019** | NR | - | - | - |  |  |  |  |
| **EVSM 2019** | NR. | - | - | - |  |  |  |  |
| **Asian Consensus 2020** | For | B | 2 | Increased Risk | ABI. |  |  | - |
|  | Against | N | 2 | No risk |  |  |  |  |

Abbreviations; **CPG**; Clinical Practice Guideline. **NICE 2012**; National Institute for Health and Care Excellence; Peripheral arterial disease: diagnosis and management Clinical guideline 147. **VASSA 2012**; Vascular Society of Southern Africa; Peripheral Arterial Disease guideline. 2012. **CEVF 2013**; Consensus Document on Intermittent Claudication from the Central European Vascular Forum (CEVF) - 3rd revision (2013). **AHA/ACC 2016**; 2016 American Heart Association/American College of Cardiology; Guideline on the Management of Patients with Lower Extremity Peripheral Artery Disease. **S3 2016**; Ärzteblatt DÄG Redaktion Deutsches. The Diagnosis and Treatment of Peripheral Arterial Vascular Disease. **ESC 2017**; 2017 ESC Guidelines on the Diagnosis and Treatment of Peripheral Arterial Diseases, in collaboration with the European Society for Vascular Surgery (ESVS). **SVS 2019**; Joint guidelines of the Society for Vascular Surgery, European Society for Vascular Surgery, and World Federation of Vascular Societies. Global vascular guidelines on the management of chronic limb-threatening ischemia. **ESVM 2019**; European Journal of Vascular Medicine. Guideline on peripheral arterial disease. 2019. **Asian Consensus**; Asia-Pacific Consensus Statement on the Management of Peripheral Artery Disease. 2020. **ABI**; Ankle-Brachial Index **NR**; No Recommendations.
\*; View full table in Appendix for parameters that suggest increased risk according to the guideline
\*\*; View Full table in the Appendix for details of recommendations suggested by the guideline

In those with no additional risk factors, the age range for screening recommendations with the more recent guideline written after 2016 (AHA/ACC, ESC, and the Asian Consensus paper) suggest screening adults over 65 years of age, while the older guidelines (VASSA and CEVF) suggest screening for those over 70 years.

The guidelines made unanimous recommendations for using ABI as the screening tool, with the older guidelines recommending further testing in the face of normal ABI in high-risk groups. Only the CEVF guideline suggested a screening interval of 2 – 3 years in high-risk groups regarding a screening interval. Risk factor Modification for High-risk groups is recommended by four guidelines.

#### Diagnostic Recommendations

The guidelines were unanimous on the decision to use ABI as the initial testing tool with predominantly strong recommendations (except VASSA, which issued a consensus recommendation). These were based on moderate level evidence, mostly except the ESC and ESVM, which utilised low-level evidence as shown in Table 5. Furthermore, the guidelines recommended further testing with methods such as Exercise A.B.I., Tp02, Pulse waveform, SPP etc., in a wide variety of circumstances, most especially when the result of the ABI is ambivalent. The recommendations were largely ungraded, and when backed with evidence, these were with low-level evidence. Notably, the NICE guideline recommends no further testing due to insufficient evidence of their utility.

**Table 5.**
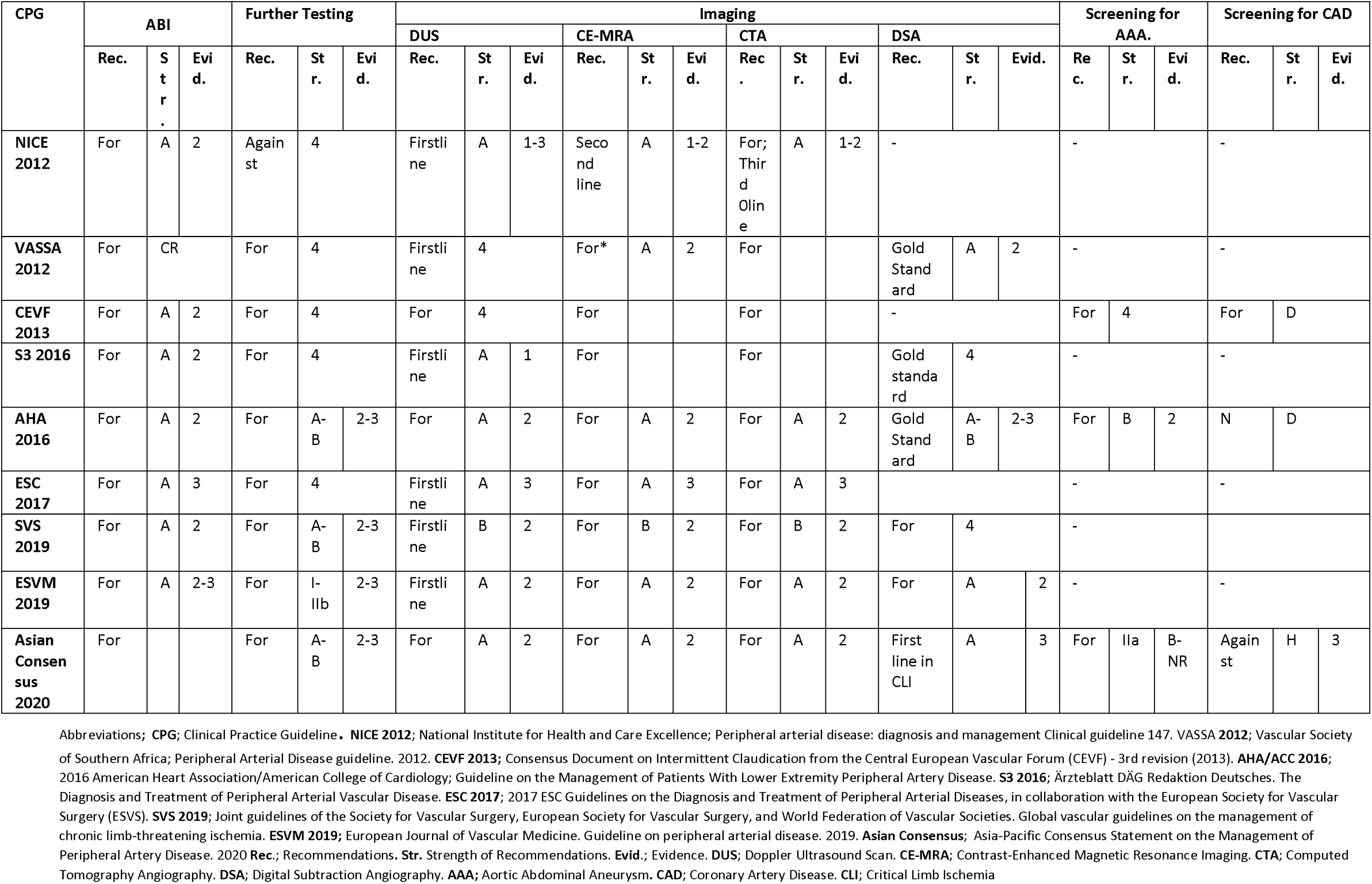
Summary of the Diagnostic Recommendations across the included guidelines.

Regarding imaging, six guidelines recommended DUS as the first-line imaging modality, with four of them making a strong recommendation. There was wide variation in the level of evidence used in making this recommendation. While CE-MRA and CTA were unanimously recommended as additional imaging, there was variation in the circumstances in which they are to be utilised. Evidence levels for the recommendations for these imaging modalities ranged between middle and low. Three guidelines noted DSA as the gold standard for imaging in PAD.. Five guidelines unanimously agreed that this modality should be reserved for cases where the arterial networks could not be adequately visualised with the other modalities.

## DISCUSSION

Overall, nine guidelines were identified and analysed in this study. In line with the study objectives, the quality of the guidelines was appraised using the AGREE tool, with the results summarised in Table 3 and Figures 1 and 2 above. This study found low scores across the applicability and stakeholder Involvement domains. The low scores in applicability can be explained by the fact that most of the guidelines did not mention monitoring or auditing criteria. Also, there was an ambiguous representation of the facilitators and barriers to implementing the guideline recommendations. Furthermore, aside from the CEVF guidelines, we observed that general practitioners, patients, and public involvement were poorly represented in the guideline development committees, resulting in low stakeholder involvement scores. This is particularly of interest, given that PAD is a largely underdiagnosed and very prevalent condition, especially among patients seen in primary care where they can and should be identified. (21) Improved General Practitioner (GP) and public involvement will improve the adoption of guideline recommendations, ultimately translating into improved patient care through early identification, which will impact a public health scale given the high prevalence of people living with PAD. A 2016 estimate placed age-standardized rates at 1930 (95%UI: 1702 to 2202) per 100,000 for women and 1658 (95%UI: 1457 to 1900) per 100,000 for men. (22) Furthermore, we noticed an improvement in the guidelines across time in all domains in our study (Figure 3), and this effect was present when we compared scores in this study to those done previously. The rigour of development scores particularly exemplifies this. The line chart in figure 3 clearly shows the rigour improving in the guidelines as they get more recent, just as observed in previous PAD guideline quality assessments. It was no surprise that we noticed better scores across the domains in this review compared with the previous studies. (6– 8) Hence we can confidently say that the PAD. guidelines are improving over time which is encouraging.

With regards to the recommendations on screening, we observed increased harmony across the guidelines of interest (over the study period) as opposed to the heterogenicity in the recommendations found in previous reviews, which included much older guidelines. Though the underlying deficiency in high-quality evidence, a randomised controlled trial (RCT) specifically designed to compare screening vs non-screening for PAD is still lacking across the guidelines, there is a general harmony in the recommendation offered to screen ‘high risk’ patients. The best evidence supporting screening comes from the VIVA study (23), where combined screening for AAA, PAD and Hypertension was offered to men aged 65 – 74 years. The PAD research community continues to anticipate an RCT to address this topic confidently. In addressing the high-risk group, there was some conflict regarding age and general silence on the contribution of gender, which is well known to influence cardiovascular risk. (24) Furthermore, in this study, we observed that just one guideline proffered a recommendation on screening intervals for PAD, which further highlights the gaps created by the absence of clear evidence.

In this paper, we also reviewed the recommendations made on the diagnosis of PAD. We found no discrepancy in utilising ABI in conjunction with clinical history and physical examination for the initial diagnosis of PAD, as solid evidence exists for this recommendation. However, there is ample evidence to show that there are occasions when ABI readings are difficult to rely on, for example, in conditions associated with hardened arteries such as diabetes. (25) In such settings, other methods were made across the guidelines for utilising such methods as Exercise ABI, TBI and T.P.P., Pulse waveforms, Oscillography and Light rheography, TcP02, amongst others. There is sparse evidence backing these recommendations with attendant variations in the circumstances in which they should be used. It is worthy to note that six guidelines strongly support the use of TBI in situations where there may be arterial hardening, such as diabetes, based on moderate level evidence. Additionally, we noticed the more recent guidelines (written after 2016) relied on weak to moderate level evidence as opposed to the older ones, which relied more on Consensus. So, while more evidence is finding its way into the guidelines clarifying this topic, we look forward to more extensive studies being conducted to enhance clarity. Furthermore, as with the recommendations on screening, these areas are of research interest to primary care physicians who are poorly represented in the PAD. guideline writing groups that could explain the apparent lack of interest in these topics.

The guidelines agreed that imaging is reserved for patients with confirmed PAD. Via initial testing methods, for whom revascularisation is being considered. The available imaging techniques suggested in the guidelines were uniform, including CFDS, CTA, CE-MRA, and DSA. It is widely acknowledged that place of practice, availability of enabling equipment, local policies and healthcare funding modalities offer some variation in the sequence/circumstances in which each modality should be chosen. For these reasons, mostly rather than based on solid evidence, the majority (six guidelines) recommended that CFDS be used as the first-line imaging of choice because it is readily available and offers the least risk to the patients (Table 5). Conversely, most of the guidelines also agreed that DSA should be reserved for cases where the arterial architecture remains ambiguous despite imaging with the other modalities due to elevated risk levels associated with its use.

And finally, with regards to screening for other arterial diseases in other vascular beds, most of the guidelines were silent. Perhaps there appears to be no additional benefit to be obtained from this. Three guidelines, CEVF, AHA and the Asian Consensus, did make recommendations. All three guidelines recommended screening for AAA via US scan, 2 of them, AHA and the Asian Consensus. The relied on evidence shows that PAD is a strong independent risk factor for AAA. However, the CEVF guideline recommends screening for CAD based on consensus recommendations. In contrast, the AHA and Asian Consensus cautioned against screening for arterial disease in other vascular beds, stating that current evidence does not justify the benefit, especially since PAD. patients should be placed on B.M.T. Current evidence has established that people living with PAD have higher rates of atherosclerotic arterial disease in other arterial beds (MI, CVA, Renal artery disease). (26) So long as there is no need for vascularisation, the treatment for all these conditions remains BMT, which includes risk factor optimisation that the PAD. patient already benefits. Justifying screening for these conditions will require evidence showing that revascularising asymptomatic forms of these diseases result in better mortality and morbidity rates, which is currently not available.

There were some obvious limitations to this study. First, this review utilised thematic qualitative analysis in synthesising guideline recommendations for comparison. Given the large volume of information contained in the guidelines, some loss of vital information was inevitable during data analysis. Extensive efforts were made to minimise these losses by utilising consistent rigorous and systematic approaches while organising the data into themes for comparation. Secondly, during the literature search for relevant CPGs, we exclusively conducted our search strategies in English. As such, it is not impossible that some relevant guidelines written during this period were not captured in this study.

## CONCLUSION

The quality of PAD. Guidelines have been improving consistently over time. Nonetheless, future guideline writers/updates should consider focusing on the guideline applicability and stakeholder involvement domains. There is less variation in screening recommendations in the recent guidelines, but a dearth of evidence persists, which could be solved with better stakeholder involvement among guideline writing committees. Finally, more research is needed to provide better evidence and thus improve guideline recommendations on imaging options for PAD.

## Data Availability

Science Data Bank

https://www.scidb.cn/anonymous/VTNleUVi

## Authors Contributorship Statement and Conflict of Interest

OU was responsible for the initial concept and design. All Authors, OU, CO, JI, OE, EC, AA, OO, participated in concept and design, extraction, analysis and interpretation of the data, critical revision of the manuscript and approval of the final version to be published.

## Appendix

1. Full Table for Summary of Screening recommendations
2. Full Table for Summary of Diagnostic recommendations
3. Search Strategies.
4. Study Protocol

