## Appendix 1 for "ORIGINAL RESEARCH; QUALITY ASSESSMENT AND COMPARATIVE ANALYSIS ON THE RECOMMENDATIONS OF CURRENT GUIDELINES ON SCREENING AND DIAGNOSIS OF PERIPHERAL ARTERIAL DISEASE; A SYSTEMATIC REVIEW"

|  | **NICE 2012** | **South Africa 2012** | **CEVF 2013** | **S3 2016** | **ACC/AHA 2016** | | **ESC 2017** | **SVS 2019** | **ESVM 2019** | **ASIAN Consensus 2020** | |
| --- | --- | --- | --- | --- | --- | --- | --- | --- | --- | --- | --- |
| **Recommendation** | - | For | For | For | For | Against | For | - | - | For | Against |
| **Strength of recommendation** | - | I | I | A | IIA | III; No Benefit | I | - | - | IIA | III; No Benefit |
| **Level of evidence** | - | B | B | I | B-NR | B-NR | C | - | - | B-NR | B-NR |
| **Target Population** | - | Patients at risk;  1. Age < 50 years with diabetes mellitus and one additional risk factor (e.g., smoking, dyslipidaemia  and hypertension)  2. Age 50 – 69 years with history of smoking and diabetes  3. Age 70 years or more  4. Leg symptoms with exertional symptoms (suggestive of claudication) or rest pain (ischaemic foot  pain)  5. Abnormal lower extremity pulse examination  6. Known atherosclerotic coronary, renal and carotid disease | In individuals whom;   1. Show arterial wall changes 2. Subjects > 70 years 3. Aged 60-69 with history of smoking or DM 4. <50 with DM +other atherosclerotic risk factors 5. >50 with metabolic syndrome | High Risk group (Not specified) | In Patients at increased risk of PAD;  1.Age >65 y  n  2.Age 50–64 y, with risk factors for atherosclerosis (e.g., diabetes mellitus, history of smoking, hyperlipidemia, hypertension) or  family history of PAD.  3.Age <50 y, with diabetes mellitus and 1 additional risk factor for atherosclerosis.  4.Individuals with known atherosclerotic disease in another vascular bed (e.g., coronary, carotid, subclavian, renal, mesenteric  artery stenosis, or AAA) | In Patients not at an increased risk of PAD | 1.Men and Women aged >65.  2.Men and Women aged <65 classified at high CV risk according to Esc guidelines  3.Men and Women >50 with family history of LEAD | - | - | Adopted AHA 2016 | In Patients not at an increased risk of PAD |
| **Screening Test** | - | ABI | ABI | ABI | ABI |  | ABI | - | - |  | ABI |
| **Further testing** | - | A more comprehensive workup of patients with PAD, considering the multiple risk factors for  atherosclerosis and the polyvascular nature of the disease | Exercise ABI can be useful if ABI is normal in at risk individuals. | - | None |  | None | - | - |  | None |
| **Intervention** | - | - known to decrease their increased risk of myocardial infarction, stroke, and death. - Smoking cessation, lipid lowering drugs, and hypertensive medication. | - Interventions known to decrease their increased risk of myocardial infarction, stroke, and death. - Smoking cessation, lipid lowering drugs, diabetes, and hypertensive medication. | - | - Statins; improves cardiovascular outcomes  - No benefit from SAPT. |  | Modification of risk factors to CVD.  No benefit from SAPT. | - | - |  | - |
| **Screening Intervals** | - | Not stated | 2-3 years | - | - | - | - | - | - |  | - |
