## Appendix 2 for "ORIGINAL RESEARCH; QUALITY ASSESSMENT AND COMPARATIVE ANALYSIS ON THE RECOMMENDATIONS OF CURRENT GUIDELINES ON SCREENING AND DIAGNOSIS OF PERIPHERAL ARTERIAL DISEASE; A SYSTEMATIC REVIEW"

| **Item** | | **NICE 2012** | | | **South Africa 2012** | | | **CEVR 2013** | | | **S3 2016** | | |
| --- | --- | --- | --- | --- | --- | --- | --- | --- | --- | --- | --- | --- | --- |
|  | | **Recommendation** | **Strength** | **Evidence** | **Recommendation** | **Strength** | **Evidence** | **Recommendation** | **Strength** | **Evidence** | **Recommendation** | **Strength** | **Evidence** |
| **Initial Testing with ABI** | | Recommended for Initial diagnosis | Strong | Moderate | For | - | - | Recommended for initial diagnosis | I | B | Recommended for initial diagnosis | A | I |
| **Further Testing for Diabetics** | | Against use of Pulse Oximetry.  Insufficient evidence for TBI and doppler wave form analysis. |  | Insufficient evidence | - |  |  | *Toe systolic pressure suggested* | - | - | TBI For diabetics with an ABI >1.3 |  | CR |
| **Other Further testing** | | - | - |  | *Exercise ABI for claudicants.*  *Toe pressure measurements and transcutaneous oxygen measurements in selected patients* |  | - | Exercise ABI for symptomatic patients with Normal ABI. | I | B | TBI and Pausatility index if ABI is implausible |  | CR |
|  |  |  |  |  |  |  |  | *Transcutaenous oxygen pressure for severe claudicants* | - | - | Oscillography and light reflection rheography in conditions like media sclerosis or acral circulatory disorders |  | CR |
|  |  |  |  |  |  |  |  |  |  |  | Stress test -Walking distance in Claudicants and for diagnosis in atypical complaints |  | CR |
| **Imaging for diagnosis of anatomical location and severity of stenosis when revascularization is considered.** | **DUS** | First - line | Strong | High - Low | First-Line |  |  | *All patients with Moderate – Severe/CLI* | - | - | First-Line | A | I |
|  | **CE-MRA** | Second - line | Strong | High - Moderate | Useful in Aorto-Iliac Disease | I | A | *For patients with severe -CLI* | - | - | Inconclusive DUS. Interdisciplinary decision with regards to therapy |  | CR |
|  | **CTA** | Third – line (If CE-MRA is not tolerated) | Strong | High - Moderate | Second-line (First line in Aorto-iliac disease) | IIa | B | *For patients with severe -CLI* | - | - | Inconclusive DUS. Interdisciplinary decision with regards to therapy |  | CR |
|  | **DSA** |  |  |  | Gold standard. Reserved for prior to surgical intervention | I | B |  |  |  | Gold standard. Inconclusive DUS. Interdisciplinary decision with regards to therapy |  | CR |
| **Screening Duplex USS scan for AAA, SAoA** |  | - |  |  |  |  |  | *Screening with Doppler of the Supra-aortic arteries and abdominal aorta* | - | - |  |  |  |
| **Screening for CAD** |  |  |  |  |  |  |  | *Always perform ECG and Echo* | - | - |  |  |  |

| **Item** |  | ACC/AHA 2013 | | | ESC 2017 | | | SVS 2019 | | |
| --- | --- | --- | --- | --- | --- | --- | --- | --- | --- | --- |
|  | | Recommendation | Strength | Evidence | Recommendation | Strength | Evidence | Recommendation | Strength | Evidence |
| **Initial Testing with ABI** | | Recommended for Initial Diagnosis. | I | B-NR | Recommended for Initial Diagnosis. | I | C | Recommended for Initial Diagnosis. | I | B |
| **Further Testing for Diabetics** | | - | - |  | - | - | - |  |  |  |
| **Other Further testing** | | TBI recommended when ABI >1.4 | I | B-NR | TBI or Doppler wave form analysis or pulse volume recordings indicated for incompressible arteries or ABI > 1.4 |  |  | TP and TBI in all patients with suspected CTLI and tissue loss | I | B |
|  |  | Exercise ABI for non-joint leg related symptoms and normal or borderline ABI + Assessing functional status | I + IIa | B-NR + B-NR |  |  |  | Consider PVR, TcPO2 or SPP when ankle or toe pressure indices cannot be assesed | II | C |
|  |  | TBI with waveform, TcPO2, SPP; For normal/borderline ABI with non-healing wounds or gangrene | IIa | B-NR |  |  |  |  |  |  |
| **Imaging for diagnosis of anatomical location and severity of stenosis when revascularization is considered.** | DUS | An option for first line; individualized decision | I | B-NR | First Line Imaging for confirmation of LEAD | I | C | First line Imaging | II | B |
|  |  |  |  |  | An option for anatomic categorization | I | C |  |  |  |
|  | CE-MRA | An option for first line; individualized decision | I | B-NR | An option for anatomic categorization | I | C | Option for second line imaging | II | B |
|  | CTA | An option for first line; individualized decision | I | B-NR | An option for anatomic categorization | I | C | Option for second line imaging | II | B |
|  | DSA (IA) | Gold standard. For confirmation when first line is inconclusive. | I | B-NR | - | - | - | Should be done for all patients with suspected CTLI | Good practice statement |  |
|  |  | First line in CLI | I | C-EO |  |  |  |  |  |  |
|  |  | Life limiting claudication with minimal response to BMT | IIa | C-EO |  |  |  |  |  |  |
|  |  | Should not be performed in asymptomatic PAD | III; Harm | B-R |  |  |  |  |  |  |
| **Screening Duplex USS scan for AAA, SAoA** |  | Is Reasonable | IIa | B-NR | - | - | - | *-* | - | - |
| **Screening for CAD** |  | *Not recommended* | - | - | - | - | - | *-* | - | - |

| **Item** |  | ESVM 2019 | | | Asian Consensus 2020 | | |
| --- | --- | --- | --- | --- | --- | --- | --- |
|  | | Recommendation | Strength | Evidence | Recommendation | Strength | Evidence |
| **Initial Testing with ABI** | | ABI as appropriate initial test | I | B - C | Recommended for Diagnosis of PAD |  |  |
| **Further Testing for Diabetics** | | In diabetes mellitus and in all those with an ABI > 1.3, toe pressure measurements and calculation of  the toe-brachial Index are recommended to detect PAD | I | B |  |  |  |
| **Other Further testing** | | Exercise ABI is useful in atypical presentations and ambiguous ABI at rest result | I | B | TBI, where available, should be measured to diagnose patients with suspected PAD when the ABI is greater than 1.40 | I | B-NR |
|  |  | In the presence of implausible ABI values, complementary methods such as TBI and calculation of pulsatility index are to be employed | II | B | Patients with exertional non-joint-related leg  symptoms and normal or borderline resting ABI  (> 0.90 and ≤ 1.40) should undergo exercise treadmill ABI testing to evaluate for PAD. | I | B-NR |
|  |  | In the case of incompressible ankle arteries, in medial calcific sclerosis, acral circulatory  disorders or ABI > 1.30, alternative methods such as the toe-brachial index, Doppler frequency analysis, oscillography or LRR or pulse volume recording  may be considered | IIb | C | In patients with normal (1.00 – 1.40) or borderline  (0.91 – 0.99) ABI in the setting of nonhealing  wounds or gangrene, it is reasonable to diagnose CLI by using TBI with waveforms, transcutaneous oxygen pressure (TcPO2), or skin perfusion pressure (SPP). | IIa | B-NR |
| **Imaging for diagnosis of anatomical location and severity of stenosis when revascularization is considered.** | DUS | Method of choice for primary diagnosis and initial evaluation of the arterial architecture | I | B | DUS, CTA, or MRA of the lower extremities is useful to assess anatomic location and severity of stenosis for patients with symptomatic PAD in whom revascularization is considered. | I | B-NR |
|  | CE-MRA | Additional diagnostic procedures –MRA, computed tomographic angiography CTA or DSA are recommended only if CCDS fails to sufficiently reveal the underlying pathology, and if proceeding to elective surgical revascularization | I | B |  |  |  |
|  | CTA |  |  |  |  |  |  |
|  | DSA (IA) |  |  |  | First line in CLI | I | C-EO |
|  |  | It is recommended that when findings are inconclusive, a second imaging method must be  applied prior to invasive procedures. Individual risk profile and the diagnostic precision of the additional method must be considered in selecting the further diagnostical procedure. | I | B | Invasive angiography is reasonable for patients  with lifestyle-limiting intermittent claudication with  an inadequate response to GDMT for whom revascularization  is being considered | IIa | C-EO |
|  |  |  |  |  | Should not be performed in asymptomatic PAD | III; Harm | B-R |
| **Screening Duplex USS scan for AAA, SAoA** |  | - |  |  | Is Reasonable | IIa | B-NR |
| **Screening for CAD** |  | *-* |  |  | Not recommended | III;Harm | C-EO |
