## Appendix 3 for "ORIGINAL RESEARCH; QUALITY ASSESSMENT AND COMPARATIVE ANALYSIS ON THE RECOMMENDATIONS OF CURRENT GUIDELINES ON SCREENING AND DIAGNOSIS OF PERIPHERAL ARTERIAL DISEASE; A SYSTEMATIC REVIEW"

Pubmed

Research Question;

Is there a significant variation in the quality and treatment recommendations of recent clinical practice guidelines on peripheral arterial disease?

Concept table

|  | P | I | C | 0 |
| --- | --- | --- | --- | --- |
|  | Adults + Elderly  Peripheral Artery Disease | Screening  Non-pharmacological  Pharmacological | None | Quality  Recommendations |
| Concepts | Peripheral Artery disease.  Chronic limb ischemia.  Critical limb ischemia. | Treatment/Management | Guideline | Recommendations  Quality |
| Free Text Terms | Arterial Disease, Peripheral. (tw)  Arterial Diseases, Peripheral. (tw)  Disease, Peripheral Arterial. (tw)  Diseases, Peripheral Arterial. (tw)  Peripheral Arterial Diseases. (tw)  Peripheral Artery Disease. (tw)  Artery Disease, Peripheral. (tw)  Artery Diseases, Peripheral. (tw)  Disease, Peripheral Artery. (tw)  Diseases, Peripheral Artery. (tw)  Peripheral Artery Diseases. (tw)  Limb  Ischemia (tw).  Chronic (tw).  Acute (tw). | Screening (tw).  Treatment (tw).  Management (tw).  Diagnosis (tw).  Pharmacological (tw) | Clinical (tw) practice (tw). Guideline (tw).  Standards (tw). | Quality (tw)  Standards (tw)  Recommendations (tw) |
| Controlled Vocabulary/MeSH | Peripheral Arterial Disease (mh)  Intermittent claudication (mh).  Lower extremity, ischemia (mh). | Diagnosis (mh).  Therapy (mh). | Practice Guidelines as Topic / standards* (mh) | Quality Improvements. (mh)  Quality Indicators. (mh) |

(((((((((((((((((((((((((((((((((Arterial Disease, Peripheral) OR (Arterial Diseases, Peripheral)) OR (Disease, Peripheral Arterial)) OR (Diseases, Peripheral Arterial)) OR (Peripheral Arterial Diseases)) OR (Peripheral Artery Disease)) OR (Artery Disease, Peripheral)) OR (Artery Diseases, Peripheral)) OR (Disease, Peripheral Artery)) OR (Diseases, Peripheral Artery)) OR (Peripheral Artery Diseases)) OR (Peripheral Arterial Disease[MeSH Terms])))) OR (intermittent claudication[MeSH Terms])) OR (limb ischemia)) AND (screening)) OR (treatment)) OR (management)) OR (diagnosis)) OR (pharmacological)) OR (Diagnosis[MeSH Terms])) OR (therapy[MeSH Terms])) AND (guidelines)) OR (guideline)) OR (standards)) OR (practice guideline[MeSH Major Topic])) AND (Quality)) OR (recommendations)) OR (quality improvements[MeSH Terms])

Example search 11/12/2020; 7014

1. Arterial Disease, Peripheral
2. Arterial Diseases, Peripheral
3. Disease, Peripheral Arterial
4. Diseases, Peripheral Arterial
5. Peripheral Arterial Diseases
6. Peripheral Artery Disease
7. Artery Disease, Peripheral
8. Artery Diseases, Peripheral
9. Disease, Peripheral Artery
10. Diseases, Peripheral Artery
11. Peripheral Artery Diseases
12. Peripheral Arterial Disease [MeSH]
13. Intermittent Claudication [MeSH]
14. Limb Ischemia
15. 1 OR 2OR 3 OR 4 OR 5 OR 6 OR 7 OR 8 OR 9 OR 10 OR 11 OR 12 OR 13 OR 14
16. Screening.
17. Treatment.
18. Management.
19. Diagnosis
20. Pharmacological
21. Diagnosis[MeSH Terms]
22. Therapy[MeSH Terms]
23. 16 OR 17 OR 18 OR 19 OR 20 OR 21 OR 22
24. Guidelines.
25. Guideline
26. Standards
27. Practice guideline[MeSH Major Topic]
28. 24 OR 25 OR 26 OR 27
29. Quality
30. Recommendations
31. Quality improvements[MeSH Terms])
32. 29 0R 30 OR 31
33. 15 AND 23 AND 28 AND 32

Search conducted on 11/12/2020. Result; 7014 references

**DETAILS OF SYSTEMATIC SEARCH STRATEGY (20/12/2020 – 30/12/2020)**

**Scopus SEARCH = 942**

TITLE-ABS-KEY ( "Arterial disease, Peripheral" OR "Arterial diseases, Peripheral" OR "Disease, Peripheral arterial" OR "Peripheral Arterial Diseases" OR "Peripheral artery disease" OR "Artery Disease, peripheral" OR "Artery Diseases, Peripheral" OR "Disease, Peripheral Artery" OR "Diseases, Peripheral Artery" OR "Peripheral Artery Diseases" OR "Limb Ischemia" OR "Chronic limb ischemia" OR "Arterial occlusive disease" OR "Artery occlusive disease" OR "Arterial occlusive diseases" OR "Artery occlusive diseases" OR "Intermittent Claudication" AND treatment OR management OR screening OR diagnosis OR therapy AND "Clinical Practice Guideline" OR "Clinical Practice Guidelines" OR guideline OR guidelines OR guide OR standard OR standards OR "Practice Guideline" OR "Practice Guidelines" OR recommendations ) AND ( LIMIT-TO ( PUBYEAR , 2021 ) OR LIMIT-TO ( PUBYEAR , 2020 ) OR LIMIT-TO ( PUBYEAR , 2019 ) OR LIMIT-TO ( PUBYEAR , 2018 ) OR LIMIT-TO ( PUBYEAR , 2017 ) OR LIMIT-TO ( PUBYEAR , 2016 ) OR LIMIT-TO ( PUBYEAR , 2015 ) OR LIMIT-TO ( PUBYEAR , 2014 ) OR LIMIT-TO ( PUBYEAR , 2013 ) OR LIMIT-TO ( PUBYEAR , 2012 ) OR LIMIT-TO ( PUBYEAR , 2011 ) OR LIMIT-TO ( PUBYEAR , 2010 ) ) AND ( LIMIT-TO ( SUBJAREA , "MEDI" ) OR LIMIT-TO ( SUBJAREA , "NURS" ) OR LIMIT-TO ( SUBJAREA , "NEUR" ) OR LIMIT-TO ( SUBJAREA , "IMMU" ) ) AND ( LIMIT-TO ( PUBSTAGE , "final" ) ) AND ( LIMIT-TO ( DOCTYPE , "ar" ) ) AND ( LIMIT-TO ( SRCTYPE , "j" ) ) AND ( EXCLUDE ( PUBYEAR , 2010 ) ) AND ( EXCLUDE ( SUBJAREA , "BIOC" ) OR EXCLUDE ( SUBJAREA , "PHAR" ) OR EXCLUDE ( SUBJAREA , "ENGI" ) OR EXCLUDE ( SUBJAREA , "IMMU" ) OR EXCLUDE ( SUBJAREA , "CENG" ) OR EXCLUDE ( SUBJAREA , "SOCI" ) OR EXCLUDE ( SUBJAREA , "ENVI" ) OR EXCLUDE ( SUBJAREA , "AGRI" ) OR EXCLUDE ( SUBJAREA , "MATE" ) OR EXCLUDE ( SUBJAREA , "BUSI" ) OR EXCLUDE ( SUBJAREA , "COMP" ) OR EXCLUDE ( SUBJAREA , "MATH" ) OR EXCLUDE ( SUBJAREA , "PHYS" ) OR EXCLUDE ( SUBJAREA , "PSYC" ) ) AND ( EXCLUDE ( EXACTKEYWORD , "Peripheral Occlusive Artery Disease" ) )

**Pubmed Search = 1723**

Search: ((((((((((((((((((((((("Peripheral arterial disease"[MeSH Terms]) OR ("Peripheral arterial disease"[Text Word])) OR ("Arterial Disease, Peripheral"[Title/Abstract])) OR ("Arterial Diseases, Peripheral"[Title/Abstract])) OR ("Disease, Peripheral Arterial"[Title/Abstract])) OR ("Diseases, Peripheral Arterial"[Title/Abstract])) OR ("Peripheral Arterial Diseases"[Title/Abstract])) OR ("Peripheral Artery Disease"[Title/Abstract])) OR ("Artery Disease, Peripheral"[Title/Abstract])) OR ("Artery Diseases, Peripheral"[Title/Abstract])) OR ("Disease, Peripheral Artery"[Title/Abstract])) OR ("Diseases, Peripheral Artery"[Title/Abstract])) OR ("Peripheral Artery Diseases"[Title/Abstract])) OR ("limb ischemia")) OR (intermittent claudication[MeSH Terms])) OR (intermittent claudication[Text Word])) AND (therapy[MeSH Subheading])) OR (therapeutics[MeSH Terms])) OR (treatment[Text Word])) OR (treatment[Text Word])) OR (disease management[MeSH Terms]) ) AND (practice guideline[Publication Type])) OR (practice guidelines as topic[MeSH Terms])) OR (clinical practice guideline) Filters: Practice Guideline, in the last 10 years, Humans, Adult: 19+ years

**TRIP database Search = 366;**

(Peripheral, Arterial, Artery, Disease, Diseases, Guidelines, Guideline) (""Peripheral arterial disease" OR "Clinical practice guidelines" OR 'Peripheral artery disease" OR "Extremity Ischemia"") from:2010 to:2020. Filter; Guideline documents.

**GUIDELINE DATABASES**

1. Guidelines International Network Library

<https://guidelines.ebmportal.com/>

Phrase Search for “Peripheral arterial disease" Found = 6

2. National guideline clearing house through Alliance for the implementation of clinical practice guidelines

<https://aicpg.org/ngc-summaries/>

Phrase Search for “Peripheral arterial disease” Found = 3

3. Canadian Medical association clinical practice guideline infobase

[https://joulecma.ca/cpg/homepage](https://joulecma.ca/cpg/homepage 4)

Phrase Search for “Peripheral arterial disease” Found = 4

4. NICE

<https://www.evidence.nhs.uk/>

Phrase Search for “Peripheral arterial disease” Found = 105

**Total citations = 3148**

| Scopus | 942 |
| --- | --- |
| Pubmed | 1723 |
| TRIP | 366 |
| Guidelines international network Library | 6 |
| National guideline clearing house through Alliance for the implementation of clinical practice guidelines | 3 |
| Canadian Medical association clinical practice guideline infobase | 2 |
| NICE | 105 |
| Targeted Internet search from expert recommendations | 1 |
| TOTAL | 3148 |

Total citations after search = 3149. Eliminate duplicates in ref manager

3112 citations left. Eliminate references written on or before 2010

3108 citations left. Elimitate references not related to PAD

333 citations left. Eliminate citations which are not clinical practice guidelines

36 citations left. Eliminate guidelines targeted at aspects of PAD management

17 citations left. Then, eliminate guidelines targeting specific patient populations

15 citations left. Repeat check for Duplicates e.g due to different languages

8 citations
